# Precision risk assessment for pediatric hospitalization using address-level data in Cincinnati, Ohio

**DOI:** 10.1101/2025.07.14.25329701

**Authors:** Carson S. Hartlage, Qing Duan, Erika Rasnick Manning, Judith W. Dexheimer, Andrew F. Beck, Cole Brokamp

**Author notes:** The authors have no conflicts of interest to disclose.

## Abstract

**Introduction:** Persistent disparities in child health highlight the need for clinical and public health research approaches to identify and address risks with greater spatial precision. This study linked residence- and neighborhood-specific socio-environmental data to population-wide healthcare data to characterize pediatric hospitalization risk for every residential address in Cincinnati, Ohio.

**Methods:** We linked hospitalization data (07/01/2016-06/30/2022) to parcel-level housing data from the Hamilton County Auditor and Cincinnati Department of Buildings & Inspections and street-range crime data from the Cincinnati Police Department. Addresses were localized to 2010 census tracts to join variables from the U.S. Census American Community Survey and Eviction Lab. Generalized random forest models estimated address-level hospitalization risk and birth-adjusted hospitalization risk, accounting for child residency using vital birth records. Model performance was assessed based on varying diagnostic thresholds; fairness was evaluated by census block-level racial demographics.

**Results:** We matched 81.5% of hospitalizations to residential addresses. Among 77,077 addresses, 7.4% had ≥1 hospitalization. Our model performed well (ROC-AUC: 0.98–0.99; PR-AUC: 0.65-0.72) in characterizing high-risk addresses, with housing violations, violent crime, and market total value among top features. The birth-adjusted model also showed high performance (ROC-AUC: 0.92-0.93; PR-AUC: 0.65-0.78) and moderate agreement with the hospitalization risk model (κ=0.43).

**Conclusions:** Our results highlight the potential of address-level modeling and multiscale data integration to build on traditional area-level analyses and advance precision population health. Future directions include geographic expansion, stakeholder engagement, and patient-level validation. This work offers a scalable approach to precisely identifying pediatric health risks, supporting targeted clinical and policy interventions.

## INTRODUCTION

Despite decades of medical advances, poor and minoritized children continue to experience adverse health outcomes at disproportionately high rates. For example, children in Hamilton County, Ohio, collectively spend ∼25,000 days in the hospital annually.^1^ If children from all area neighborhoods were hospitalized at the same rate as those from the most affluent quartile of neighborhoods, hospitalization days could decrease by >30%.^1^ These inequities have lasting effects, as early exposure to adverse socioenvironmental conditions can negatively impact health and well-being into adulthood.^2^ Moreover, hospitalizations impose burdens on patients, families, and healthcare systems.^3,4^

The integration of medical, social, and environmental data can prompt a more complete understanding of how and why child health varies between and within communities and inform better strategies to address these disparities. Traditional population health approaches often rely on area-level (e.g., ZIP Code, neighborhood, census tract) characteristics. Area-based approaches, however, can be limited by shifting and ill-defined boundaries, infrequent data updates, inherent sample-based uncertainty, and the assumption of homogeneity among all individuals within the unit of area.^5^ Further, deploying interventions at a neighborhood scale can be resource intensive and impractical.

Multiscale linkages are required to utilize large amounts of extant medical, social, and environmental data available at different spatiotemporal resolutions and extents. Integrated assessments of residence- and neighborhood-based influences can enhance public health interventions and the provision of patient care but remain uncommon in population health and clinical practice.

Addresses within the electronic health record (EHR) provide an opportunity for linkages to potentially meaningful datasets, enabling more granular studies of place-based health determinants.^6^ However, addresses, including those in the EHR, can be messy and unstructured, and traditional geocoding approaches are often not precise enough for address-level matching.^6–8^ As such, address-level data integration is technically challenging and may be limited by poor match rates. Moreover, relevant previous work is limited in scope and has not tackled a comprehensive population study.^9–11^ Local municipalities often make address-based datasets publicly available, including information on legal units of land such as residential properties, enabling linkage of data on property type, condition, value, age, and more. This offers significantly greater spatial precision than area-based approaches, as a single census tract may contain several hundred residential properties and addresses (Figure 1).

**Figure 1.**
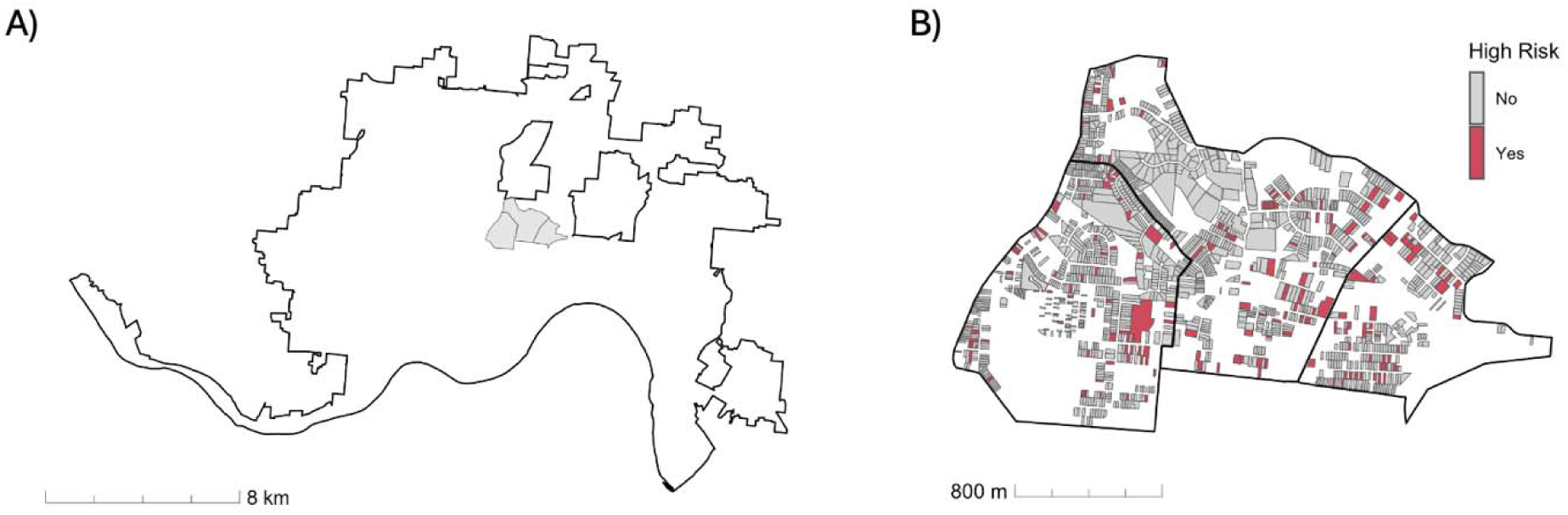
A map of census tract and residential property geographies in Cincinnati, Ohio. A) Three census tracts located directly northeast of Cincinnati Children’s within the city of Cincinnati. According to 2019 American Community Survey 5-year averages, these tracts are comprised of a total population of 9,656 people in 4,173 households, including 3,055 children across 1,148 households. B) Geographies of 1,582 residential properties representing 1,799 addresses within the same three census tracts. Properties associated with high-risk addresses as classified by the hospitalization risk model at the top 7.4% diagnostic threshold are colored red.

Increased precision can come at the cost of data sparsity; hospitalizations are rare at the individual- and address-level.^12^ Machine learning models are well-suited to handle this challenge, along with the collinearity and complex interactions present in many socio-environmental datasets. This makes such models valuable for integrating diverse contextual information and translating it into actionable insights for addressing health inequities.^13^ Despite the availability and granularity of address-level data, there remains a lack of comprehensive models integrating residence- and neighborhood-specific data with population-wide health outcomes that can inform targeted population health and clinical practices.

We sought to develop a machine learning model that integrates population-wide healthcare data with residence- and neighborhood-specific variables to identify addresses associated with high pediatric hospitalization risk. Our dataset included 77,077 residential addresses linked to 10,085 total pediatric hospitalizations occurring during our 6-year study period and 30 input features obtained from open data sources. Using this approach, we generated address-level risk for child hospitalization (ARCH) scores for every address in Cincinnati.

## MATERIALS AND METHODS

This work was completed as part of the Responding to Identified Sociomedical Risks with Effective Unified Purpose (RISEUP) Study, approved by the Cincinnati Children’s Institutional Review Board. The data we used for temporal validation was covered under a second protocol, also approved by the Cincinnati Children’s Institutional Review Board.

### Study setting and population

This study took place in Cincinnati, Ohio, a dense urban area that is home to Cincinnati Children’s Hospital Medical Center (CCHMC), the region’s only pediatric hospital, and about 65,000 children and adolescents. CCHMC handles approximately 95% of all pediatric hospitalizations in Hamilton County, and coverage is likely higher within city limits, which are fully contained within the county.

The study population was defined in October 2024 as all residential addresses in the Cincinnati Area Geographic Information System (CAGIS).^14^ CAGIS supports permitting, licensing, inspections, code enforcement, planning, zoning, and asset management by disseminating information on parcels, legally recognized areas of land. Addresses associated with parcels with land use types presumed to be non-residential (e.g., park, vacant or commercial lot) and those without a land use type were excluded.

### Address-level hospitalization

We used the RISEUP cohort, which consists of individuals under 18 years of age from Hamilton County, Ohio, who were hospitalized at CCHMC between July 1, 2016, and June 30, 2022.

We extracted patient addresses from the EHR and linked them to the address-based dataset of parcel identifiers obtained from CAGIS using the addr package (0.4.0) in R.^15^ The purpose of addr is to clean, parse, and match addresses to a reference dataset using a natural language processing (NLP) machine learning model. Addresses associated with parcels located outside Cincinnati’s boundaries were filtered using parcel centroid latitude and longitude and the city boundaries maintained by the City of Cincinnati.^16^ Addresses assumed to not represent patients’ long-term residences (e.g., Hamilton County Job and Family Services, CCHMC) were excluded. For each address, we calculated our primary outcome by summing the total number of hospitalizations during the study period.

To account for potential confounding by the number of children living at each address, a secondary outcome was modeled incorporating birth records as a proxy for child residency. Birth data between July 1, 2016, and June 30, 2022, were obtained from the Ohio Bureau of Vital Statistics and matched to addresses using the method described above. Birth-adjusted hospitalization scores were calculated as the number of hospitalizations minus the number of births, normalizing scores based on the likelihood that children resided at each address. For example, the birth-adjusted model would understand a single-family home with five hospitalizations and a large apartment building with five hospitalizations as having different magnitudes of risk. Because most addresses had zero births, we chose to subtract rather than divide birth counts to avoid input scores of Infinity (e.g., one hospitalization divided by zero births) or undefined (e.g., zero hospitalizations divided by zero births).

### Input Feature Data

We harmonized 30 input features for each address from public data sources: 11 at the residence level, linked via parcels or custom buffered summaries of point- or grid-based geospatial data, and 19 at the neighborhood level, linked via census tract geographies (Table 1).

**Table 1.**
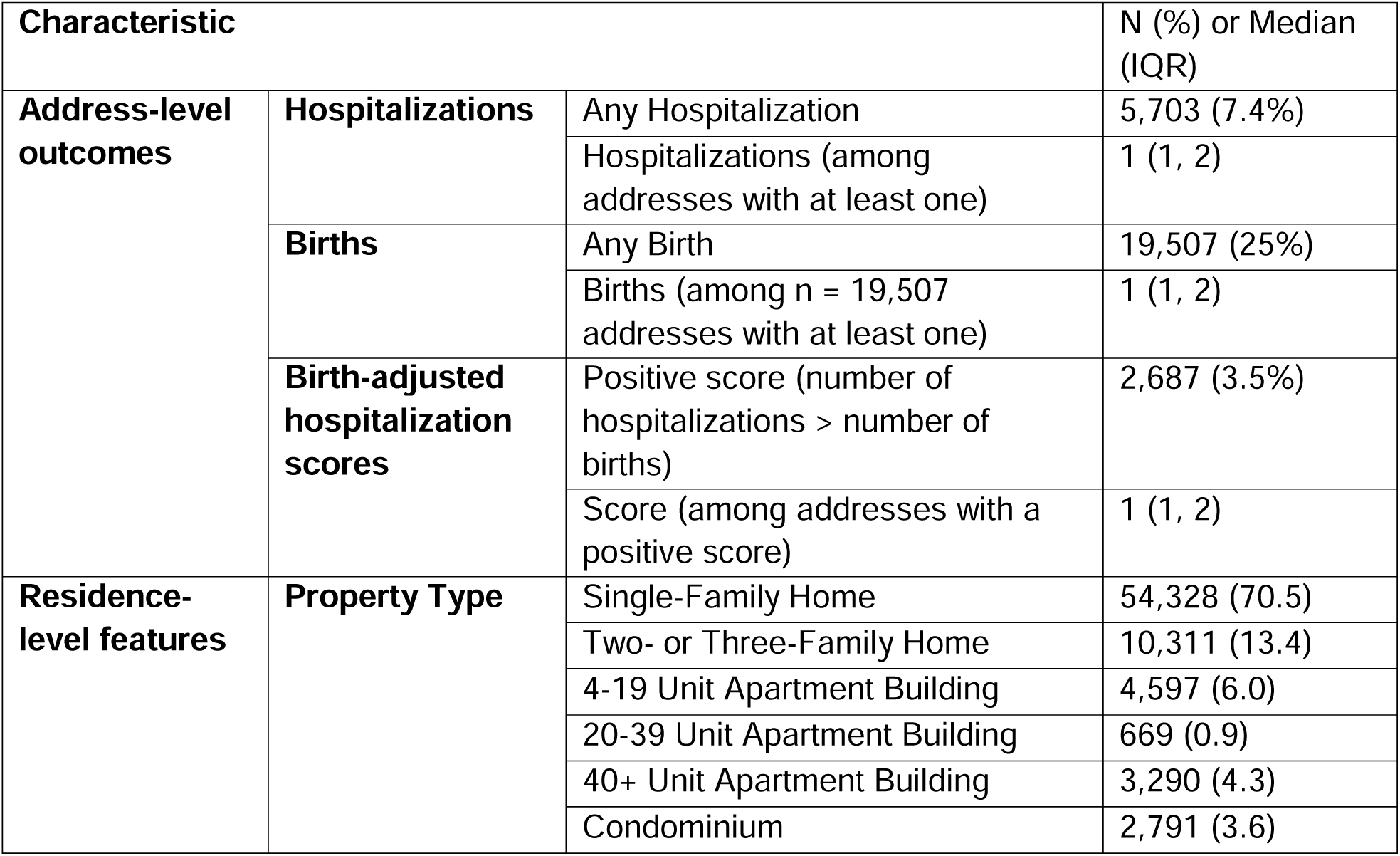
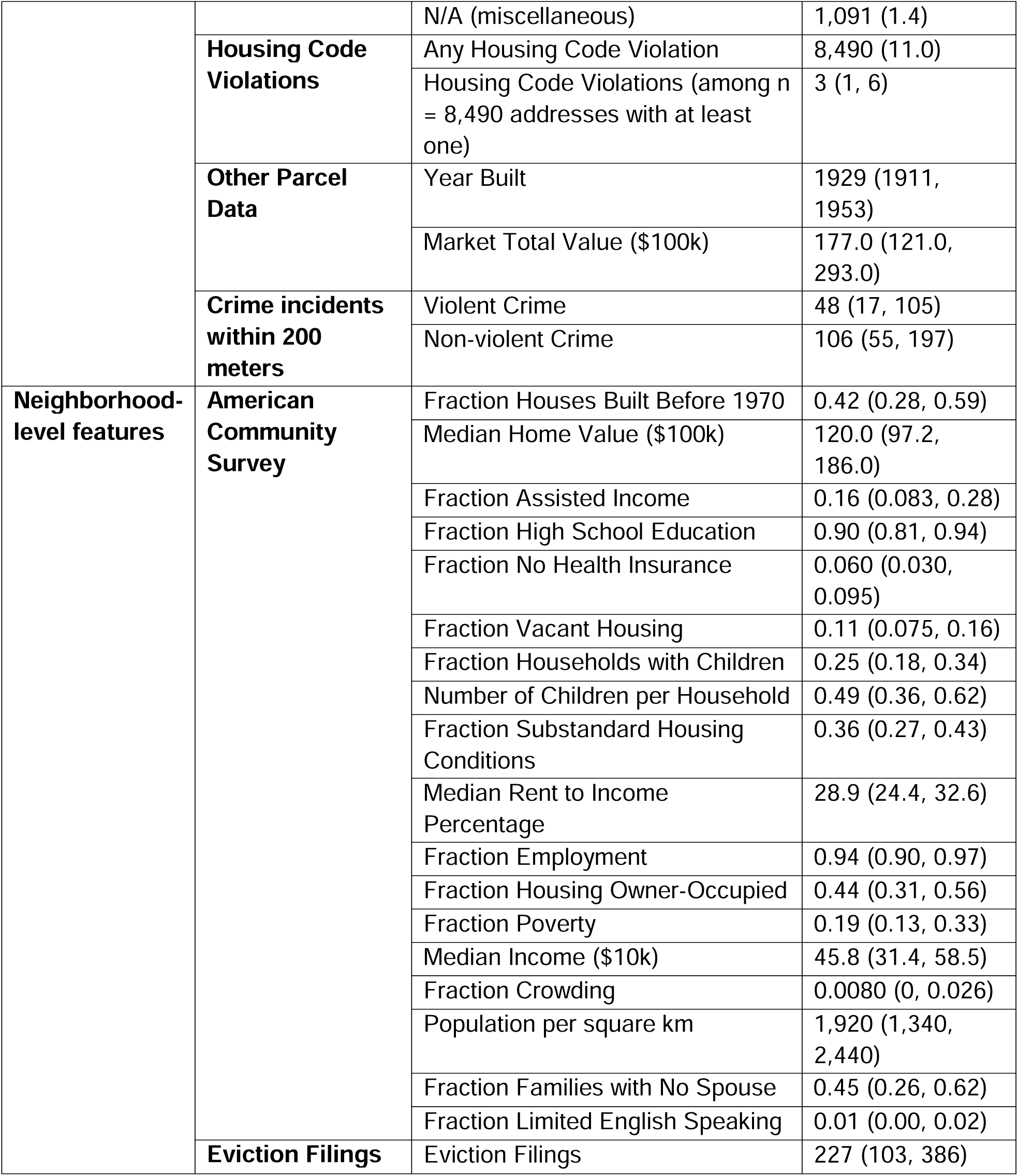
Characteristics of 77,077 addresses included in the study. IQR: Interquartile range.

#### Residence-level features

Data on housing code violations by parcel identifier were obtained from the City of Cincinnati Department of Buildings & Inspections.^17^ We summed the total number of health-related (e.g., deteriorated paint, mold, pest infestation) enforced housing code violations between July 1, 2016, and June 30, 2022, for each parcel. Housing code violations in Cincinnati are complaint-driven; there are no proactive housing inspections. Other parcel data, including year built, market total value, and property type, were retrieved from CAGIS and the Hamilton County Auditor.^14,18^ These data are updated at least every three years, with the last update occurring in January 2024. Property types were operationalized as shown in Table 1. Condominiums included the labels “condominium unit”, “landominium”, and “condo or pud garage.” Separate variables were not included for less common, miscellaneous property types (e.g., “other residential structure”, “metropolitan housing authority”, “mobile home / trailer park”; n=1,091 addresses).

Crime data were obtained from the Cincinnati Police Department via Open Data Cincinnati.^19^ These data include daily crime incidents localized to street ranges (e.g., 32XX Burnet Ave). We tabulated the number of incidents characterized in the dataset as “violent” occurring between July 1, 2016, and June 30, 2022. We summarized this number using the crime street ranges overlapping with a 200-meter (approximately 1/8-mile) radius of the parcel centroid latitude and longitude. This radius was selected based on the size of a typical city block. Incidents labelled “property” or “other” were summarized similarly as non-violent crime incidents.

For addresses matched to multiple parcels (n=522), parcel data were summarized as follows: median of market total value, year built, violent crime incidents, and non-violent crime incidents; mean number of housing code violations. Some addresses were missing data on year built (n=465) and market total value (n=127). To eliminate outliers due to data entry errors, market total values below the 1^st^ percentile and above the 99^th^ percentile were considered missing (n=1,210).

#### Neighborhood-level features

Addresses were localized to 2010 census tract geographies by intersecting parcel centroids with tract boundary polygons and joined to U.S. American Community Survey 2019 5-year average data. Four census tracts were missing median home value data (n=1,025).

Data on eviction filings were obtained from the Eviction Lab, which began releasing tract-level data in January 2020.^20^ Monthly eviction filing counts from January 2020 through June 2024 were summed. At the start of the COVID-19 pandemic, Hamilton County suspended municipal court proceedings between March 19, 2020, and June 1, 2020, and filings dropped. Our study period includes this time, and we assumed that variation among tracts did not significantly differ during periods impacted by COVID-19. Eviction data were interpolated from 2020 to 2010 census tract geographies using census block-level population weights.

### Model development

We used generalized random forest models. This approach uses an adaptive nearest-neighbor method where predictions are made based on weighted averages of outcomes from data points that are “neighbors” of a target observation.^21^ These local neighborhoods are adaptively defined by the structure of bagged decision trees, which partition the feature space to prioritize predictive accuracy. Random forests dynamically tailor the size and composition of local neighborhoods for each prediction, allowing them to effectively capture complex relationships in data. Because generalized random forest handles missingness as an attribute, we did not impute missing data. Regression forest models were fit using the grf package (2.3.2) in R with 1000 trees, a sample fraction of 0.5, minimum node size of 5, and mtry of 26.

### Model evaluation and calibration

Model outputs were defined as ARCH scores. Performance metrics, including area under the receiving operator curve (ROC-AUC), area under the precision-recall curve (PR-AUC), sensitivity, specificity, positive predictive value (PPV), and negative predictive value (NPV), were calculated based on classification of ranked ARCH scores at representative thresholds. For the hospitalization risk model, we assessed performance for two outcomes: addresses with at least one hospitalization and addresses with at least two hospitalizations (top 7.4% and 2.4% of addresses, respectively). For the birth-adjusted model, we assessed performance for addresses with more hospitalizations than births (top 3.5%) and the top 2.4% of addresses. We evaluated the agreement between the two models using Cohen’s kappa for the top 2.4% outcome. We also used calibration plots to visualize the relationship between input and output scores.

Fairness was evaluated by assessing model performance across four subgroups defined by census block-level racial composition. Addresses were localized to census blocks by intersecting parcel centroids with 2020 census block geographies and joined to 2020 U.S. Census data. They were then stratified into quartiles based on the proportion of the population in the census block identifying as white alone. Addresses missing block-level demographic data (n=83) were excluded. Model performance fairness was evaluated after applying the optimal cutoff as defined by Youden’s index. We used two fairness metrics: equalized odds, the maximum difference in sensitivity or specificity between subgroups; and equal opportunity, the maximum difference in sensitivity between subgroups.

Temporal robustness for the hospitalization risk model was assessed using address-level hospital admissions between July 1, 2022, and June 30, 2023. We re-calibrated thresholds to match the hospitalization prevalence of the validation dataset and calculated the same performance metrics described above; we also did this analysis using raw address-level hospitalization counts from the training data instead of ARCH scores. We were unable to temporally validate the birth-adjusted model because we do not have access to updated vital records data.

### Model interpretation

The most important variables were determined by calculating importance as a weighted sum of how many times trees split on each feature up to a maximum depth of 4 splits. To supplement variables of importance by examining variable interactions within the random forest models, shallow decision trees were fit to the model output scores. The trees were verified by fitting decision trees to the corresponding input scores. The trees allowed us to visualize the structure and stratify ARCH scores. We also utilized partial dependence plots to visualize the relationship between market total value and risk, stratified by property type. All model interpretations are intended to be descriptive.

## RESULTS

We matched 81.5% of hospital admissions to residential addresses in Cincinnati, resulting in 10,085 hospital admission encounters linked to 5,704 unique addresses. Of 77,077 total addresses, 5% had exactly one hospitalization, and 2.4% had multiple hospitalizations. In Table 1, we characterize addresses based on our input features. Most addresses are single-family homes (70.5%), with smaller proportions of multi-family homes (13.4%) and apartment buildings (11.2%). Approximately 11% of addresses had at least one housing code violation, with a median of three violations (IQR: 1, 6) among those affected. Within a 200-meter radius of each address, the median number of violent crime incidents was 48 (IQR: 17, 105), and the median number of non-violent incidents was 106 (IQR: 55, 197).

Our hospitalization risk model produced AUC values of 0.99 and 0.98 in identifying the top 2.4% and 7.4% highest-risk addresses, respectively (Table 2). PPVs were 0.34 and 0.56, respectively (Table 2). The ten most important variables were split between residence- and neighborhood-level variables, with housing violations and violent crime as the most important (Figure 2).

**Figure 2.**
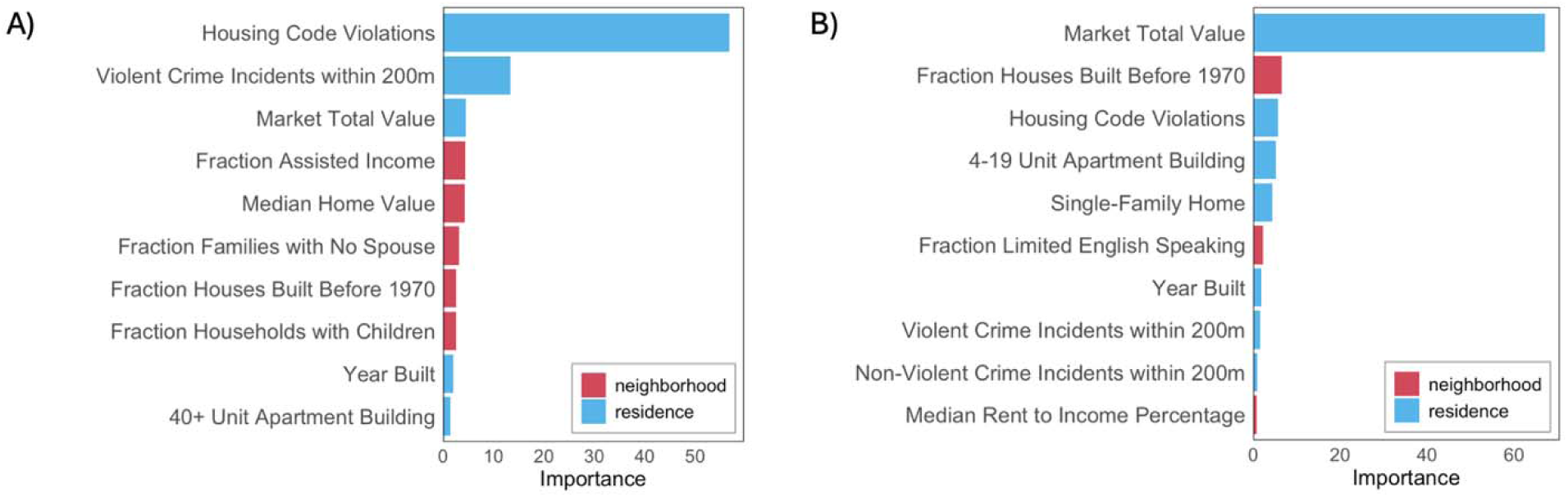
Variable importance plots. A) Top 10 most important features in the hospitalization risk model. B) Top 10 most important features in the birth-adjusted hospitalization risk model.

**Table 2.** Performance metrics for both hospitalization risk models for two outcomes. ROC-AUC: area under the receiving operator curve; PR-AUC: area under the precision-recall curve; CI: confidence interval; PPV: positive predictive value; NPV: negative predictive value.

| Model Outcome | Threshold | ROC-AUC (95% CI) | PR-AUC (95% CI) | Sens | Spec | PPV | NPV |
| --- | --- | --- | --- | --- | --- | --- | --- |
| hospitalization | Multiple hospitalizations (top 2.4%) | 0.99 (0.99-0.99) | 0.65 (0.63-0.68) | 0.97 | 0.95 | 0.34 | 0.99 |
|  | At least one hospitalization (top 7.4%) | 0.98 (0.98-0.98) | 0.72 (0.71-0.73) | 0.94 | 0.96 | 0.56 | 0.99 |
| birth-adjusted hospitalization | Top 2.4% | 0.93 (0.92-0.95) | 0.68 (0.65-0.72) | 0.89 | 0.97 | 0.26 | 0.99 |
|  | Hospitalizations > Births (top 3.5%) | 0.92 (0.91-0.93) | 0.65 (0.63-0.68) | 0.85 | 0.96 | 0.42 | 0.99 |

Calibration plots assessed the agreement between risk scores and actual hospitalization risk, and visual inspection revealed approximately linear relationships (Figure S1; Figure S2). For the hospitalization risk model, the median number of hospitalizations for addresses with scores above 1 was 3 greater than addresses with scores of 0 or below. Similarly, for the birth-adjusted model, the median birth-adjusted hospitalization for addresses with scores above 0.5 was 4 greater than addresses with scores below -0.5.

Birth records were linked to 19,507 addresses, or 25% of all addresses. Among these, 42.7% (n=8,129) had multiple births during the study period. Performance of the birth-adjusted model remained robust, with AUC values of 0.93 and 0.92 for the top 2.4% and top 3.5%, respectively (Table 2). The birth adjustment resulted in different rankings of the most important features (Figure 2).

We evaluated the temporal robustness of the hospitalization risk model by linking scores to address-level hospitalizations one year after the training data, resulting in 1,534 hospital admissions at 1,050 addresses (1.4% of all addresses). Performance remained fair with an ROC-AUC of 0.75 (95% CI: [0.74, 0.77]) for detecting addresses with at least one hospitalization (Table S1). Using raw hospitalization counts from the training data to forecast in lieu of ARCH scores demonstrated a slightly lower ROC-AUC of 0.68 (95% CI: [0.66, 0.69]; Table S2).

Market total value, housing code violations, violent crime, year built, and fraction of houses built before 1970 were among the ten most important variables for both models (Figure 2). The hospitalization and birth-adjusted hospitalization risk models differed in which addresses were classified as high-risk, indicating sensitivity to outcome definition. Assessing agreement between the two models for the top 2.4% threshold produced a Cohen’s kappa of 0.427, indicating moderate agreement. The property type distribution among addresses classified as high-risk by both models resembled that of the overall population (70% single-family homes; Table 1). In contrast, only 30% of addresses characterized as high-risk by the hospitalization risk model alone were single-family homes, compared to 86% for those identified solely by the birth-adjusted model. Decisions trees fit on ARCH scores provide visualization of the most influential features and their interactions (Figure 3; Figure 4). By splitting on key residence- and neighborhood-level variables, the trees demonstrate how distinct pathways contribute to high hospitalization risk and can support interpretation of model results. Fraction of households with children, violent crime incidents, and market total value appeared in both trees, and the top split points were housing code violations and 40+ unit apartment building for the hospitalization risk model and birth-adjusted model, respectively (Figure 3; Figure 4). Partial dependence plots of market total value by property type showed a positive relationship between value and hospitalization risk and a negative relationship between value and birth-adjusted hospitalization risk, with differences between property types (Figure S3; Figure S4).

**Figure 3.**
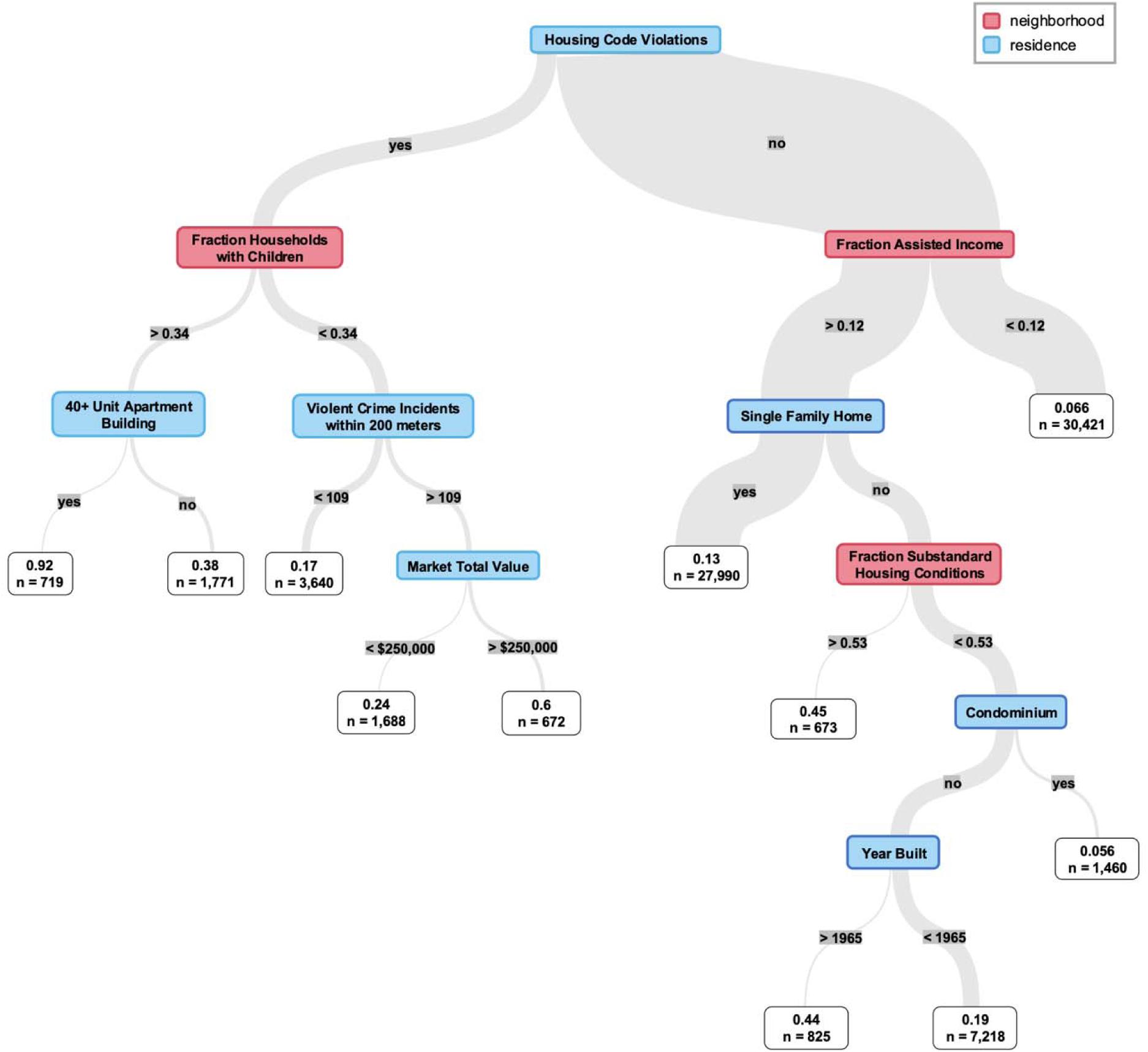
Decision tree fit to the output scores of the hospitalization risk model.

**Figure 4.**
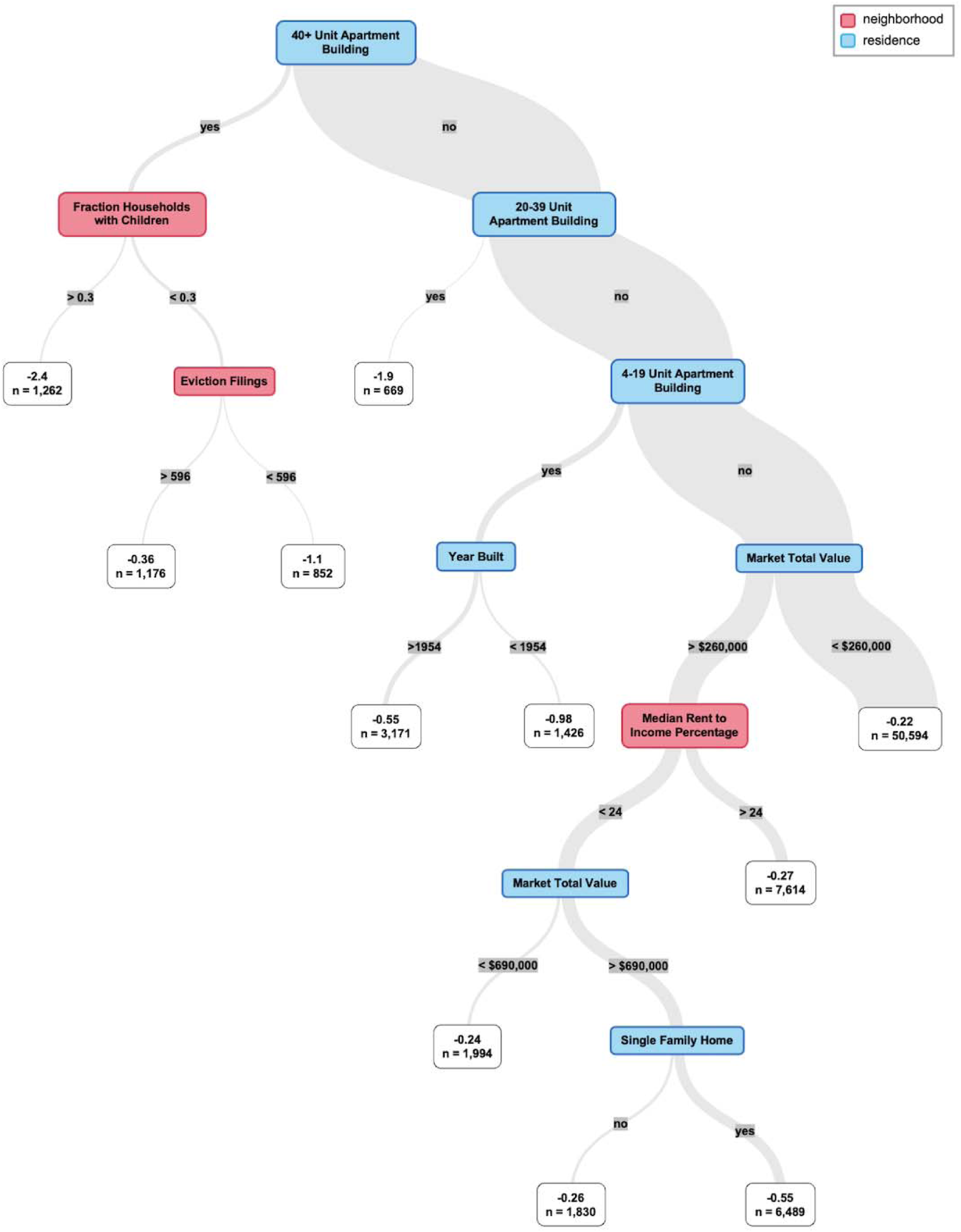
Decision tree fit to the output scores of the birth-adjusted hospitalization risk model.

Avondale is a neighborhood with one of the highest hospitalization rates in the city.^1^ The tracts shown in Figure 1 are contained in Avondale, and tract-level proportion of addresses classified as high-risk by the hospitalization risk model at the top 7.4% threshold ranged from 11.0% to 18.4%.

Our fairness assessment demonstrated some difference in performance by census block-level racial composition. The highest metric, indicating the greatest disparity between groups, was for the birth-adjusted model at the top 2.4% outcome, where equalized odds and equal opportunity highlight that sensitivity differed by 0.14 between the first quartile (greatest proportion of white residents) and the fourth quartile (lowest proportion of white residents; Table 3). There were differences in property type distribution among quartiles; the proportion of addresses that are single-family homes in the first, second, third, and fourth quartiles was 78.1%, 70.5%, 72.1%, and 60.5%, respectively.

**Table 3.** Fairness metrics for hospitalization and birth-adjusted hospitalization risk models based on subgroups defined by census block-level racial demographics.

| Model Outcome | Diagnostic Threshold | Equalized Odds | Equal Opportunity |
| --- | --- | --- | --- |
| hospitalization | Multiple hospitalizations (top 2.4%) | 0.10 | 0.04 |
|  | At least 1 hospitalization (top 7.4%) | 0.09 | 0.06 |
| birth-adjusted hospitalization | Top 2.4% | 0.14 | 0.14 |
|  | Hospitalizations > births (top 3.5%) | 0.09 | 0.09 |

## DISCUSSION

We developed machine learning models to estimate child hospitalization risk for every residential address in Cincinnati, leveraging open residence- and neighborhood-level data linked to population-wide healthcare data. We operationalized ARCH scores using ranked percentile thresholds, and our approach demonstrated strong performance in identifying addresses at high-risk for pediatric hospitalization, both total and birth-adjusted.

While neighborhood-level studies of health risk “hot spots” are well established, the use of parcel-level data in population health research is still emerging. Previous studies have used parcel-level property value data to evaluate the impact of socioeconomic status on obesity and cancer screening.^22–24^ Parcel-level lead service line data have also been used to examine racial disparities in lead exposure.^25^ Several studies have leveraged parcel data to study links between housing and health. Parcel- and building-level indicators of housing quality, including housing code violations, have been linked to asthma-related emergency department use, COVID-19 infection risk, and claims data for housing-sensitive health conditions.^26–28^ One study used a combination of parcel, custom location buffer, and census tract data to predict home pest and mold exposures for children with asthma.^29^

Healthcare research leveraging address-based data linkage is also limited. Geocoding approaches can result in poor match rates, so direct address matching often requires NLP methods.^6–8^ Other studies have used NLP-based address matching to identify patients residing in long-term care homes or other healthcare-associated settings, to measure residential mobility from the EHR, as well as for linkage across eviction, Medicaid, and homeless shelter records.^9–11,30,31^ Our group has demonstrated this approach in Hamilton County, Ohio, to estimate lead exposure risk, leveraging both parcel data and address matching.^32^ We build on this work here by implementing parcel data and address matching at a population- and city-wide scale, linking EHR data across pediatric conditions.

While other clinical machine learning models have incorporated area-level geomarkers data into patient-level predictions,^13^ we use residential addresses as the unit of study. This has several advantages, including the ability to isolate housing-specific factors, emphasizing residence and neighborhood characteristics over individual clinical or demographic attributes. The prominence of residence-level features in our variables of importance graphs (Figure 2) and decision trees (Figure 3; Figure 4) also underscores the value of this approach. In addition, it is privacy-preserving, as ARCH scores do not link back to any individual’s protected health information (since many addresses share the same score) and demonstrate better forecasting performance than historical hospitalization counts alone. This allows us to share granular health insights with interested partners without compromising patient privacy. Finally, address-level risk identification provides a foundation for precision population health strategies. Precision population health is an emerging field that aims to deliver “the right intervention to the right population at the right time,” leveraging accurate and timely data; ARCH scores are well-suited to these approaches.

ARCH scores can drive primary and secondary prevention efforts in both clinical and policy spheres. In the clinic, they could be used to inform referral to medical-legal partnerships^33^ or other housing advocates and targeted screening and guidance for housing-related health conditions and environmental exposures. In the policy sphere, insights from our model could drive targeted housing inspection for code enforcement and litigation against landlords with substandard housing. Partnerships with municipalities, such as the City of Cincinnati, could leverage these insights to allocate resources more effectively in ways aimed at reducing housing-related health disparities. Community organizations focused on neighborhood safety, housing quality, and tenant rights may also be interested in address-level health insights for advocacy and resource management. By linking address-level health insights to actionable interventions, our framework offers a comprehensive approach to mitigating risk factors and potentially reducing hospitalization rates.

We sought to account for child population distribution by modeling a birth-adjusted outcome, which normalized scores by the likelihood of children residing at each address. By using birth records, we assumed that the characteristics associated with a child being born at an address are the same as those associated with a child living at an address. This model demonstrated similar performance but changed the importance of input features and the classification of many high-risk addresses. These differences highlight that many features may reflect both the propensity for an address to be the residence of a child and the propensity for child hospitalization. The two models could be leveraged for specific use cases; for example, using ARCH scores to target addresses across the entire city for intervention and using birth-adjusted ARCH scores to assess individual-level risk for a child with a known address.

While novel, our approach has important limitations. Our study focused exclusively on Cincinnati, limiting generalizability. However, the internal validity of our models is strengthened by the near-complete population coverage (>95%) of pediatric hospitalization events and full population coverage of residential addresses. Also, while the model itself is not directly translatable to other populations or areas, we provide a framework for other researchers to adapt this approach to their population and data availability.

Although our primary goal was capturing cumulative risk rather than prediction into the future, we used one year of more recent hospital admissions data (1 year after training data) to assess the validity and temporal robustness of ARCH scores. As expected, performance metrics dropped but remained fair. ARCH scores also demonstrated better forecasting ability than previous hospitalization counts alone, highlighting the contribution of residence- and neighborhood-level features to predicting future hospitalization. Further work would be needed to investigate potential model overfitting and/or temporal drift in addresses that experience hospital admissions before using ARCH scores as a forecasting tool.

Our birth-adjusted model aimed to isolate characteristics driving high hospitalization risk by adjusting for the number of children living at each address. However, our approach had shortcomings, as property type appeared to still be an important feature in this model (Figure 2; Figure 4). Residential mobility is a major confounder of the capability of birth records to represent child population distribution. Alternative approaches could leverage EHR encounters data or commercial data sources to better estimate the number of children associated with each address over the study period.

Historical inequities in housing and healthcare make fairness a critical consideration for deployment of ARCH scores. While algorithmic fairness literature has borrowed a 0.2 threshold from legal precedent of disparate impact in employment,^34^ the 0.14 difference in sensitivity seen in the birth-adjusted model for the top 2.4% outcome is still concerning. In general, model performance was lower for the quartile of addresses with the lowest census block-level white population. It is critical that ARCH scores do not contribute to further neglect of historically disinvested communities and the children who live there. Fairness issues may be driven by several factors, including disparities in property type composition among racial demographic-based subgroups (17.6% difference between first and fourth quartiles) and bias inherent in our data sources.^35^ Utilizing publicly available data, even at a high spatial resolution, does not capture dynamic, context-specific information, and parcel data lack detailed information on individual units. This is particularly salient for multi-family homes, where data validity may be lower than for one child living in a single-family home, leading to potential exposure misclassification. Similarly, using vital records to infer child residence may be associated with similar biases. For housing code violations data, complaint-driven systems have been shown to be biased, and violations may be unequally enforced across neighborhoods.^36,37^ Finally, using block-level demographic data for fairness analysis has limitations, as it may not be reflective of individual residences, families, or children, and scores should be interpreted in context, with consideration of accuracy-fairness tradeoffs. Fairness mitigation approaches, including debiasing and post-calibration adjustment, should be explored in the future.

Future work should expand to include the rest of Hamilton County to allow for insights into a larger population. Expansion both within our area and to other areas around the country will require more data harmonization and increased open data efforts, and smaller municipalities may not have the technical infrastructure for data sharing. For example, crime and housing code violations data are largely unavailable in areas of Hamilton County outside of Cincinnati. In addition, greater incorporation of time-varying and historical data would improve model robustness and enable evaluation of policy impacts and evolving risk patterns, rather than relying solely on cross-sectional snapshots to estimate average characteristics over a historical period. Further, our processes, including the use of open data, can be specialized to other geographic areas through model retraining and adaptation to the data available in other municipalities.

Multi-sector collaboration and the integration of multi-dimensional data from diverse sources will be vital to the scalability and portability of efforts. For example, commercial providers such as Regrid (regrid.com) offer access to nationwide harmonized parcel data, and initiatives such as the Ohio Geographically Referenced Information Program (OGRIP) and MassGIS seek to make statewide parcel data publicly available.^38,39^ Adding features tailored to specific health conditions could refine the model’s utility for targeted interventions. For instance, incorporating greenspace or air pollution measures could be used for respiratory-related hospitalizations, while proximity to airports or major highways can inform lead poisoning risk assessments. Other potential enhancements include identifying rental properties, including linkage to landlords, and integrating address-level data on evictions or property renovations, which can be indicators of housing instability and in-home environmental hazards. The model could be further specialized to assess risk at rental properties or in specific neighborhoods. Additionally, children in foster care, a high-risk population, were likely underrepresented in our dataset due to address filtering that excluded hospitalization records associated with Hamilton County Job and Family Services (HCJFS), limiting our ability to fully assess their risk. Future work could include partnership with HCJFS to more accurately link admissions to foster children’s addresses of residence. Further, this modeling framework could be applied to ED visits, preventive care completion, or total cost of care as potential outcomes alongside hospitalizations. While this would require different interpretation, it could provide valuable insights into broader patterns of healthcare utilization.

Further model development, calibration, and communication should incorporate feedback from partners and end-users to ensure that outputs are actionable, interpretable, and trustworthy. Operationalization of outputs and supporting interpretability will be critical to integrating this tool into real-world settings. Finally, validation at the patient level should be assessed with a prospective healthcare cohort to evaluate overall performance and model fairness.

By integrating open address-level data with hospital utilization records in a machine learning model, we have demonstrated the potential of a novel framework for identifying health risks with high spatial precision without compromising patient privacy.

## Data Availability

All data for predictor variables are available online at the links provided. We are required to protect the individual-level privacy of healthcare and birth records. They cannot be shared.

https://github.com/geomarker-io/parcel

https://github.com/geomarker-io/addr

https://github.com/carson-hartlage/ARCH

https://github.com/geomarker-io/hh_acs_measures

https://github.com/geomarker-io/xx_address

## ACKNOWLEDGEMENTS

Births per address data used in this study were obtained from the Bureau of Vital Statistics, Ohio Department of Health (ODH). Use of these data does not imply ODH agrees or disagrees with any presentations, analyses, interpretations, or conclusions.

