## Supplemental Figures and Tables for "Precision risk assessment for pediatric hospitalization using address-level data in Cincinnati, Ohio"

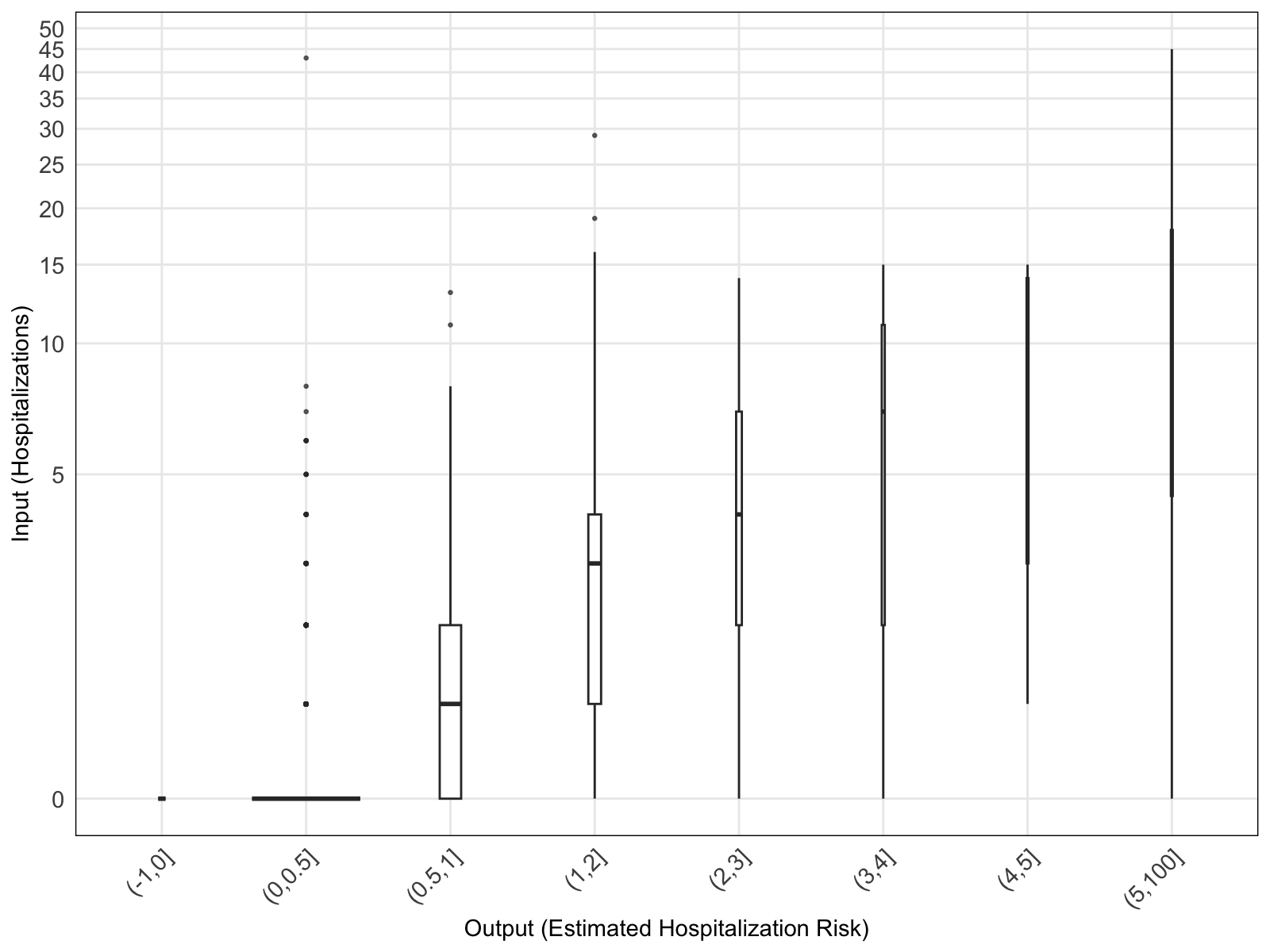
**Figure S1. Calibration plot for the hospitalization risk model.** The width of the boxes is proportional to the square root of the number of addresses per bin.

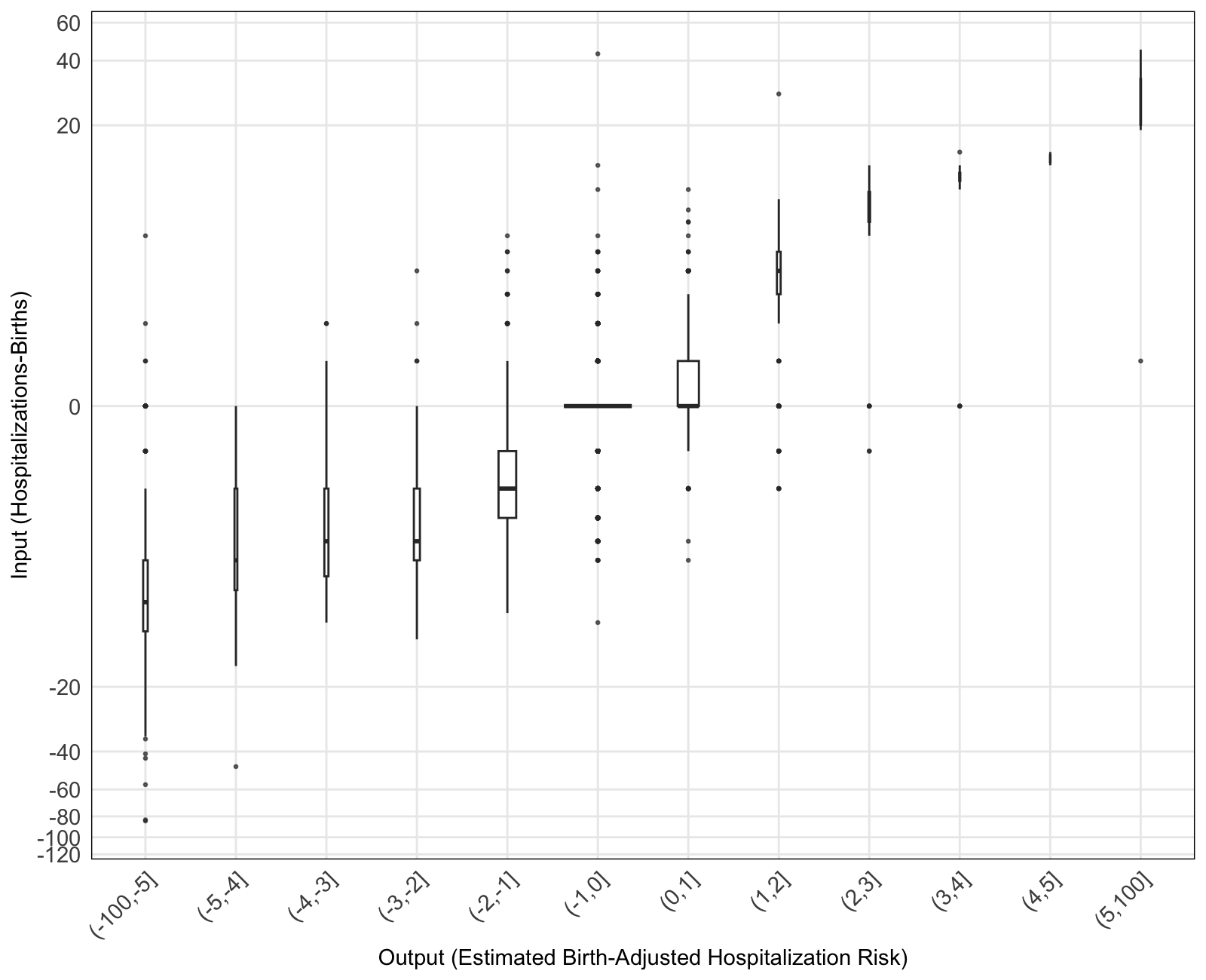
**Figure S2. Calibration plot for the birth-adjusted hospitalization risk model.** The width of the boxes is proportional to the square root of the number of addresses per bin.

**Table S1. Performance metrics for temporal validation (one year after training data) of the hospitalization risk model.** ROC-AUC: area under the receiving operator curve; PR-AUC: area under the precision-recall curve; CI: confidence interval; PPV: positive predictive value; NPV: negative predictive value.

| **Model Outcome** | **Threshold** | **ROC-AUC (95% CI)** | **PR-AUC (95% CI)** | **Sens** | **Spec** | **PPV** | **NPV** |
| --- | --- | --- | --- | --- | --- | --- | --- |
| hospitalization | Multiple hospitalizations (top 0.3%) | 0.75 (0.74-0.77) | 0.06 (0.04-0.09) | 0.53 | 0.86 | 0.03 | 0.99 |
|  | At least one hospitalization (top 1.4%) | 0.75 (0.74-0.77) | 0.09 (0.08-0.11) | 0.53 | 0.86 | 0.03 | 0.99 |

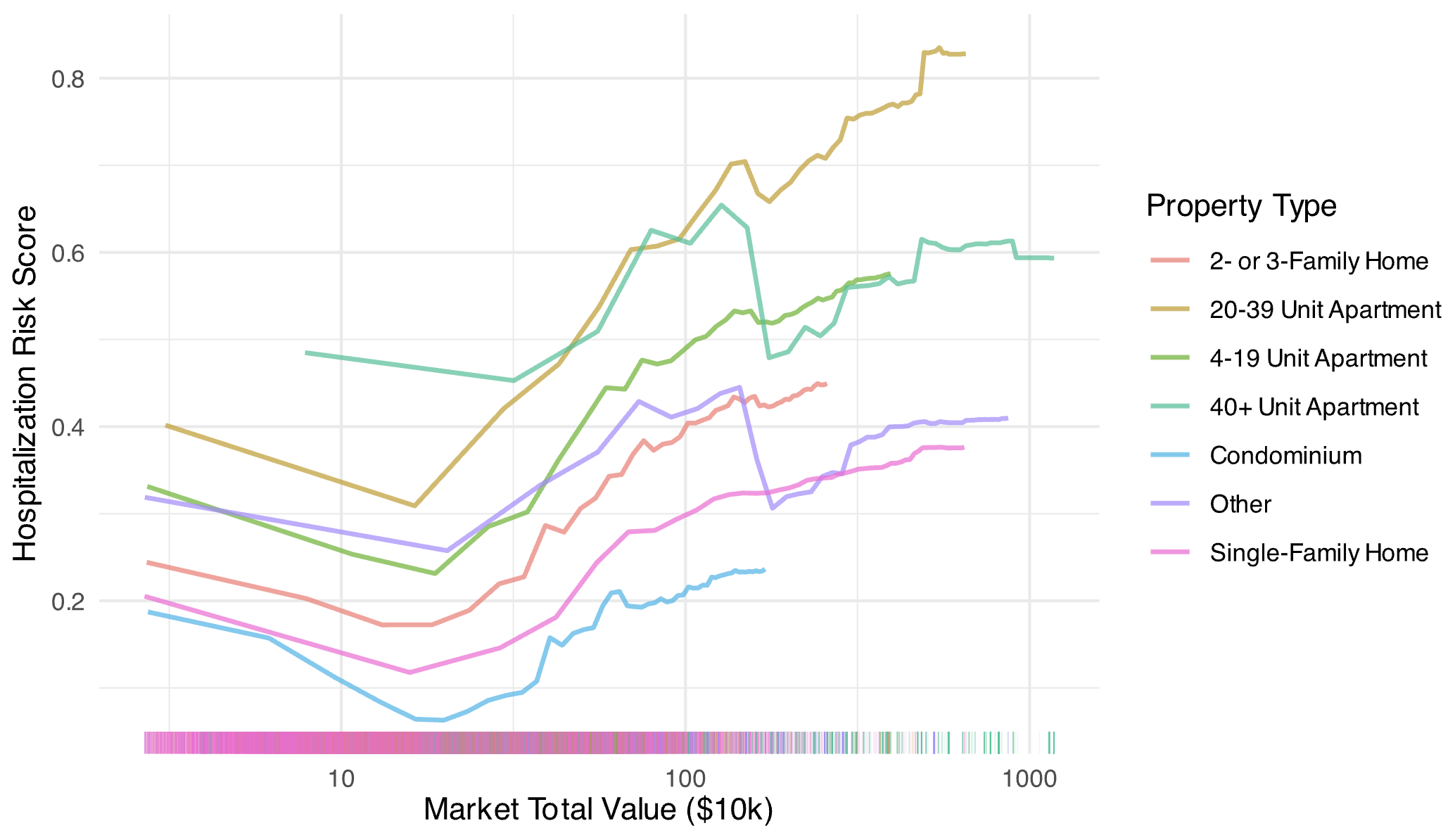
**Figure S3. Partial dependence plot of market total value by property type for the hospitalization risk model.**

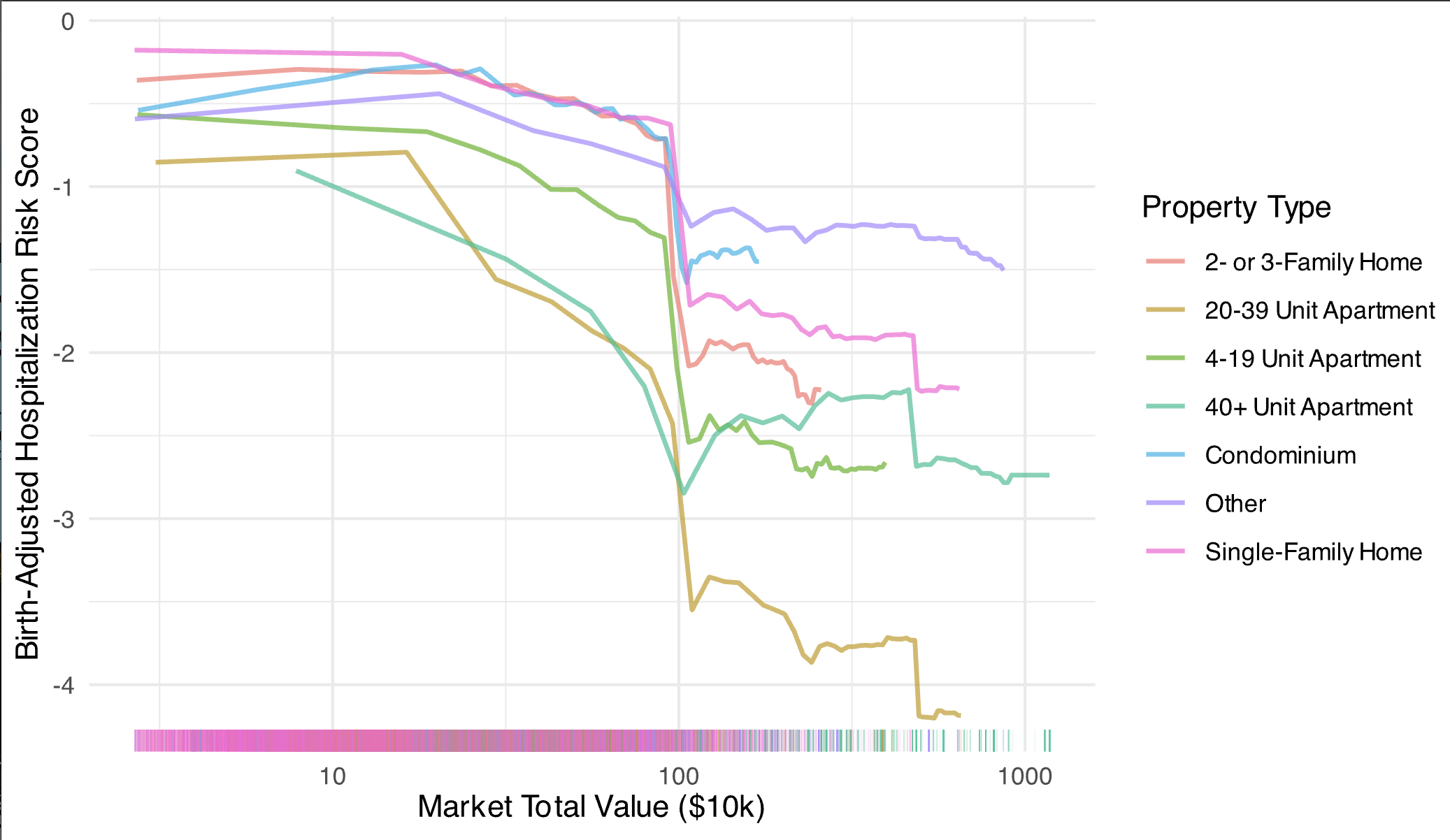
**Figure S4. Partial dependence plot of market total value by property type for the birth-adjusted hospitalization risk model.**
